# Home-based supervised cardiorespiratory interval training decreases post-stroke fatigue and improves cardiorespiratory fitness—an RCT

**DOI:** 10.1101/2025.11.12.25340123

**Authors:** Anna Bråndal, Maria Svedjebrant, Ylva Nilsagård, Per Wester

## Abstract

**Introduction:** Post-stroke fatigue (PSF) affects nearly half of all stroke survivors and significantly hinders rehabilitation and daily functioning. There is no established treatment. Low cardiorespiratory fitness may contribute to PSF, suggesting aerobic training as a potential intervention.

**Methods:** In this two-center, randomized, open-label, blinded-endpoint trial, we evaluated a home-based supervised cardiorespiratory interval training program (HS-CITP) in individuals with PSF (Swedish Fatigue Assessment Scale [S-FAS] ≥28) 1–7 months post-stroke. Participants were randomized (1:1) to either HS-CITP or usual care with self-directed activity following early supported discharge. The intervention included three weekly cycling sessions at 70%–80% of maximum heart rate over eight weeks. The primary outcome was self-reported fatigue (S-FAS); the secondary outcome was peak oxygen uptake (VO₂peak, mL/kg/min) post-intervention.

**Results:** Forty-five participants were randomized; 43 completed follow-up (HS-CITP: n=22; control: n=21). Adherence to HS-CITP was 92%, with no adverse events. Compared with the control group, HS-CITP significantly reduced fatigue (mean between-group difference −5.35 S-FAS points; 95% CI −9.03 to −3.67; p<.001) and improved cardiorespiratory fitness (+4.48 VO₂peak mL/kg/min; 95% CI 3.41–5.54; p<.001).

**Conclusion:** Supervised home-based interval training significantly reduced PSF and improved fitness, with high adherence and no safety concerns. These findings support integrating structured aerobic exercise into stroke rehabilitation. Larger, longer-term trials are needed to confirm the durability of this benefit.

**Registration:** URL: https://www.clinicaltrials.gov; Unique identifier: NCT03458884

## Introduction

Stroke is the second leading cause of death and disability worldwide and a contributor to disability-adjusted life years among neurological disorders (1). Post-stroke fatigue (PSF) affects about 50% of survivors and is consistently reported as one of the most challenging long-term symptoms. (2). Even in mild cases, PSF limits physical activity, perpetuating deconditioning and functional decline (3).

PSF is multifactorial; low levels of cardiorespiratory fitness is a key a contributor (4). Meta-analyses show a moderate association between reduced fitness and higher fatigue levels, with depression and anxiety often confounding this relationship (4, 5). Cardiorespiratory fitness, commonly measured as peak oxygen consumption (VO₂peak), reflects the capacity for sustained dynamic exercise (6). VO₂peak is markedly reduced after stroke, with values ranging from 26% to 87% of age- and sex-adjusted norms (7). This deficit increases the energy cost of daily activates potentially worsening fatigue (8, 9).

Despite guideline recommendations for aerobic exercise (10), PSF remains underexplored as a treatment target. Evidence for non-pharmacological interventions is limited (11, 12), and implementation of structured aerobic training faces logistical barriers (13). Exercise may improve fitness, enhance blood flow, and promote neuroplasticity (14, 15, 16), yet few trials have evaluated its effect on PSF (17). High-intensity interval training (HIIT) offers superior gains in aerobic capacity compared to moderate-intensity training and appears feasible and safe post-stroke (17, 18. 19).

Reducing PSF is a top research priority (3, 20), but no randomized controlled trial has compared structured aerobic exercise with standard care using fatigue as the primary outcome (5). This study aimed to determine whether a home-based, supervised cardiorespiratory interval training program (HS-CITP) added to early supported discharge reduces PSF and improves VO₂peak compared with usual care. We hypothesised that HS-CITP would lower fatigue and enhance cardiorespiratory fitness.

## Methods

The study was approved by the Swedish Ethical Review Authority (Umeå Dnr 2015-420-3M and Gävle Dnr 2019-03207). The reporting of this study was adhered to CONSORT guidelines (21). The study was registered at clinicaltrials.gov before the inclusion of participants (NCT03458884).

### Data availability

The data set analyzed during this study is available from the corresponding author upon reasonable request

### Study design and setting

This study was a 1:1 prospective randomized open-label trial with blinded evaluators (PROBE-design) (22), conducted at two sites.

The study was conducted in collaboration between the Stroke Centre, Umeå University hospital, and the ESD-team in Gästrikland. Two physiotherapists specialized in neurology (A.B. and M.S.) supervised the intervention, which was delivered in participants’ homes. Participants inclusion began in September 2018 at the Umeå site and expanded to include the Gästrikland site in September 2019. National and local COVID-19 restrictions caused one interruption of patient inclusion at the Umeå site (total time, 6 months) and two interruptions of inclusion at the Gävleborg site (total time, 12 months).

### Participants

Inclusions criteria were adult stroke survivors aged >18 years, diagnosed with either ischemic or hemorrhagic stroke, living independently in Umeå or Gästrikland, who were at least 1 month but not more 7 months post-stroke at the time of enrolment. Further inclusion criteria were self-rated PSF ≥28 according to the Swedish Fatigue Assessment Scale (S-FAS) (23) and being physically able to participate in cardiorespiratory interval training using an ergometer cycle. Exclusion criteria were severe stroke (i.e., modified Rankin Scale >3) (24), unstable pulmonary or cardiac disease, serious co-morbidity, and severe cognitive dysfunction measured with Montreal Cognitive Assessment (MOCA) ˂ 26 (25).

Before entering the study, participants had received stroke unit care followed by Early Supported Discharge (ESD) service at Stroke Centre, Umeå University hospital or from ESD-team in Gästrikland. The ESD service (26) provides support and rehabilitation for patients in their own environment. The rehabilitation includes training in ADL (e.g., dressing, cooking, walking), health information, psychological support, and coordination with services like homecare. Information about PSF, including support and practical guidance on how to identify and manage fatigue symptoms in daily activities (activity adaptation, prioritization, balancing physical activity and rest) was included as a part of the ESD service at the study sites.

### Procedure and randomization

All participants received both oral and written information about the study from the study personnel at each site (A.B. and M.S.). Individuals who met the inclusion criteria, agreed to participate, and provided written informed consent underwent medical examination by a stroke physician, followed by pre-treatment assessments of fatigue (S-FAS) and cardiorespiratory fitness (VO₂peak) in a physiological test laboratory prior to randomization. After completing pre-treatment assessments, participants were randomized to either control group (receiving standard ESD service) or intervention group (receiving S-CITP add on standard ESD service). Randomization was stratified by sex, age, and stroke severity (Modified Rankin Scale 0–1 vs. ≥2) using the web-based program Minim (27). Minimization was applied to achieve balanced group allocation upon study entry with respect to key significant variables.

### Intervention with Home-based, Supervised Cardiorespiratory Interval Training Program (HS-CITP)

Participants allocated to the intervention group preformed HS-CIPT sessions on an ergometer cycle for 30–40 minutes, three times per week for eight weeks. Each session consisted of a 10-minute warm-up at 50% of peak heart rate, followed by four 4-minute high-intensity intervals at 70%–80% of peak heart rate, interspersed with 3-minute periods of active recovery, and concluded with a 5-minute of cool-down. During each 4-minute interval, participants rated their perceived exertion every minute using Borg Rating of Perceived Exertion (RPE) scale (28) every minute during the 4-minute intervals. All training sessions were supervised and guided face-to face by an experienced physiotherapist (A.B. or M.S.).

Heart rate was continuously monitored, and individual training ranges (70%–80% of maximal heart rate) were determined from baseline cardiorespiratory fitness testing. Participants were instructed and encouraged to exercise within this range and to reach an exertion level (heavy) corresponding to 15–16 on the Borg Rating of Perceived Exertion (RPE) scale.

### Control treatment (standard ESD service)

Participants in the control group received standard ESD service, which included information about PSF, as well as support and practical advice on how to identify and manage fatigue symptoms in daily tasks, such in the adaptation and prioritization tasks, engaging in physical activity, and incorporating recovery. They were also encouraged to stay physically active.

### Outcome measures

All outcome assessments were conducted in the same order at each testing time point and test site. Clinical outcomes were assessed before randomization (pre-treatment test), and 1–2 weeks after the intervention period (post-treatment test).

The primary outcome was measured using S-FAS (23). The S-FAS is a 10-item self-report scale that evaluates fatigue symptoms using a 5-point Likert scale ranging from 1 (never) to 5 (always). Scores for items 4 and 10 are reversed (i.e., 1=5, 2=4, 3=3, 4=2, 5=1) before calculating the total score. The overall score is the sum of all items (including the recoded scores for item 4 and 10) and ranges from 10–50, with higher scores indicating more severe fatigue. The S-FAS is valid, reliable, recommended, and has been previously tested in stroke patients (23).

The physiotherapists delivering the intervention were aware that the participants had scored above 28 points on the pre-treatment S-FAS. The post-treatment S-FAS was completed independently by the participants and submitted to a research nurse at the Stroke Center, Umeå University hospital, who was blinded to participants’ group allocation. The S-FAS post-treatment rating remained unknown to those involved in delivering the intervention until the study was completed.

An incremental cardiopulmonary exercise test (29) performed on an ergometer cycle was used to measure the secondary outcome, cardiorespiratory fitness (VO₂peak). The exercise test on the partisipants was conducted by staff at the Umeå Movement and Exercise Laboratory (UMEX) at Umeå University and the Lugnet Sports Science Institute (LIVI) laboratory, Falun at Dalarna University. The test evaluators were blinded to participant group allocation. The equipment used for the exercise testing included a respiratory mass spectrometer (AMIS 2015; Innovision A/S, Odense, Denmark, Gas concentration measurement, Oxigraf, Inc, Mountain View, CA, USA), an ergometer bike (Monark 839E, Exercise AB, Sweden), and a heart rate monitor (Polar V800, Polar Electro Inc, Finland). The exercise test commenced at 30 watts, with increments of 10 watts every 60 seconds until exhaustion. Participants rated their perceived exertion on the Borg RPE scale (28) every minute during the test. The test was discontinued at a Borg RPE rating of 17 (very hard) or a respiratory exchange ratio of 1.0, based on medical considerations and to prevent undue physiological stress in an inexperienced and apprehensive individual.

### Statistical analysis

The sample size was determined to detect a between-group mean difference of 9 points on the S-FAS (23), assuming a common standard deviation of 10 points, a two-sided α =0.05, and 80% power. Using a two-sample t-test with equal allocation, the required sample size was 20 participants per group (40 total). To allow for potential dropout, we planned to enrol 50 participants (25 per group). Analyses were conducted according to the intention-to-treat principle.

Baseline data were summarized using mean, standard deviation, and range for continuous variables, and frequencies and percentages for categorical variables. Univariate analyses of covariance (ANCOVA) were conducted to examine the effects on the intervention on the primary outcome, PSF and the secondary outcome, cardiorespiratory fitness (VO2peak), while statistically controlling for age, sex, functional status (modified Rankin scale), and baseline (pre-test) values for each respective outcome. Group (intervention or control) was included as a fixed factor in both models. The dependent variables were S-FAS scores and post cardiorespiratory fitness, expressed in milliliters of oxygen per kilogram body weight per minute (mL/kg/min). Assumption of normality, homogeneity of variance, linearity, and homogeneity of regression slopes were tested and met for both ANCOVA models. Statistical analyses were performed using IBM SPSS Statistics, version 29.

## Results

Between 1 September 2018 and 31 December 2025, 50 participants met the inclusion criteria. A total of 45 participants were randomized, with 23 allocated to the intervention group and 22 to the control group. One participant in each group withdrew after randomization and baseline testing (Figure 1) Baseline characteristics were comparable between groups at the pre-treatment assessment (Table 1). Adherence to the training program was high in the intervention group. Among the 22 participants who completed intervention, the mean number of completed training sessions was 22 out of a planned 24 (SD =1.6; range 20–24), corresponding to an average adherence rate of 92%. All participants completed at least 83% of the prescribed sessions. The intervention was carried out without any reported adverse advents or safety concerns from either participants or the physiotherapists. After 8 weeks of HS-CIPT, the intervention group reported significantly lower fatigue scores compared to the control group at post-treatment, with a mean difference of 5.35 points (p <.001; 95% CI 3.67–9.03) (Figure 2). The intervention group also demonstrated a significantly greater improvement in cardiorespiratory fitness, with an increase in VO₂peak of 4.48 mL/kg/min compared to the control group (p < .001; 95% CI 3.41, 5.54) (Figure 3).

**Figure 1.**
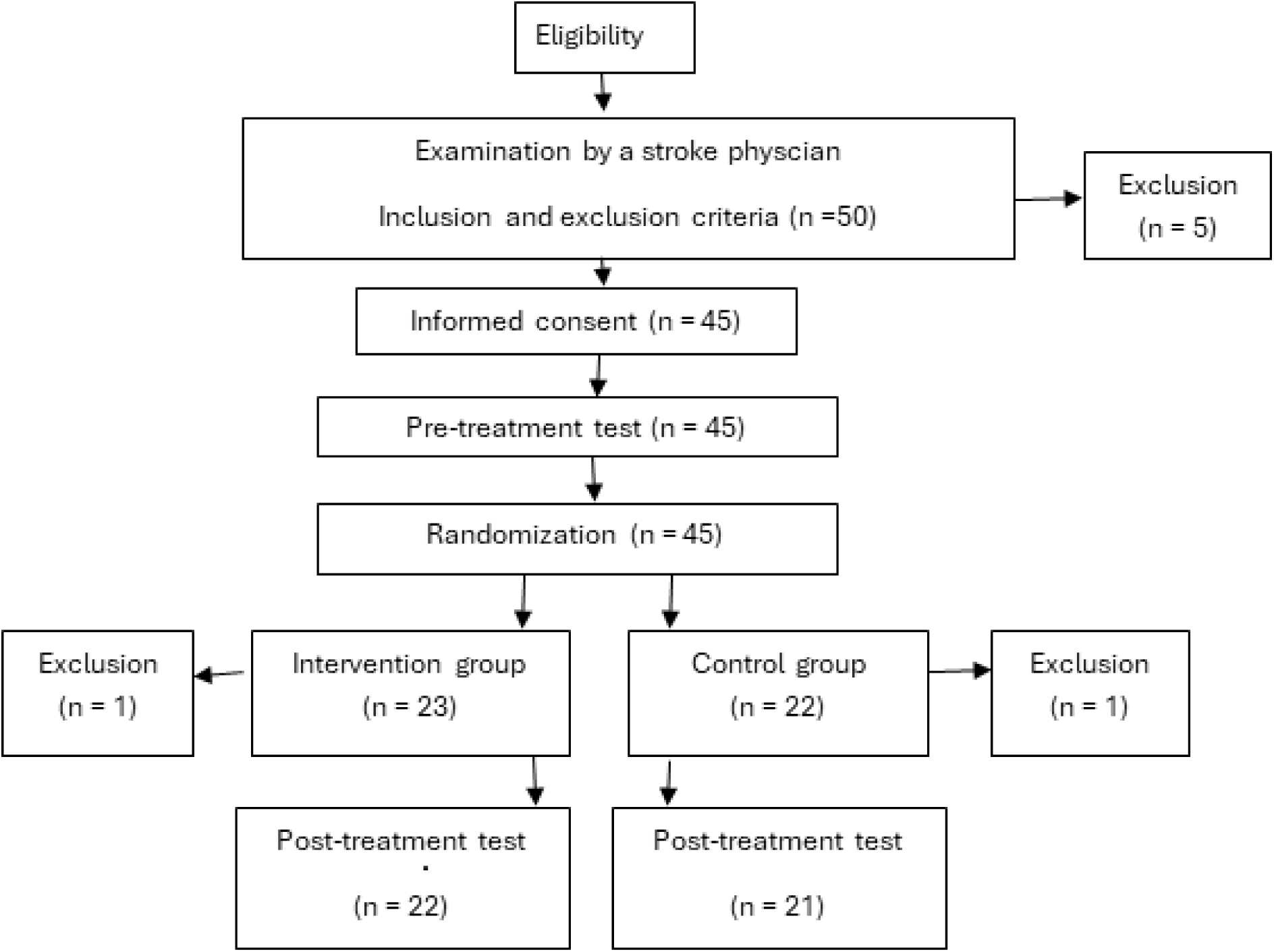
Flowchart of study procedures

**Figure 2.**
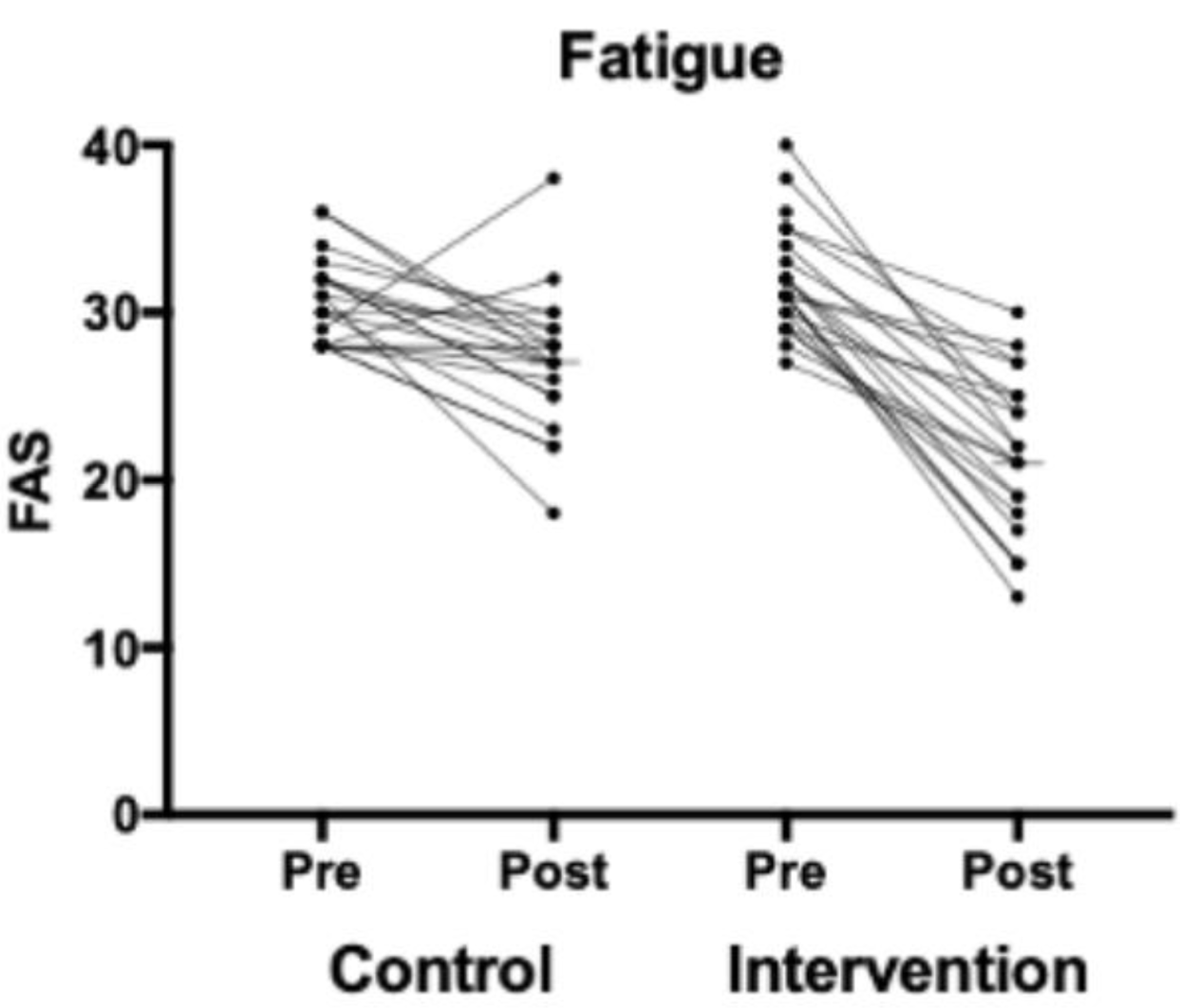
Results for post-stroke fatigue measured with the Swedish Fatigue Assessment scale (S-FAS) in the control and the intervention groups at pre-and post-treatment tests

**Figure 3.**
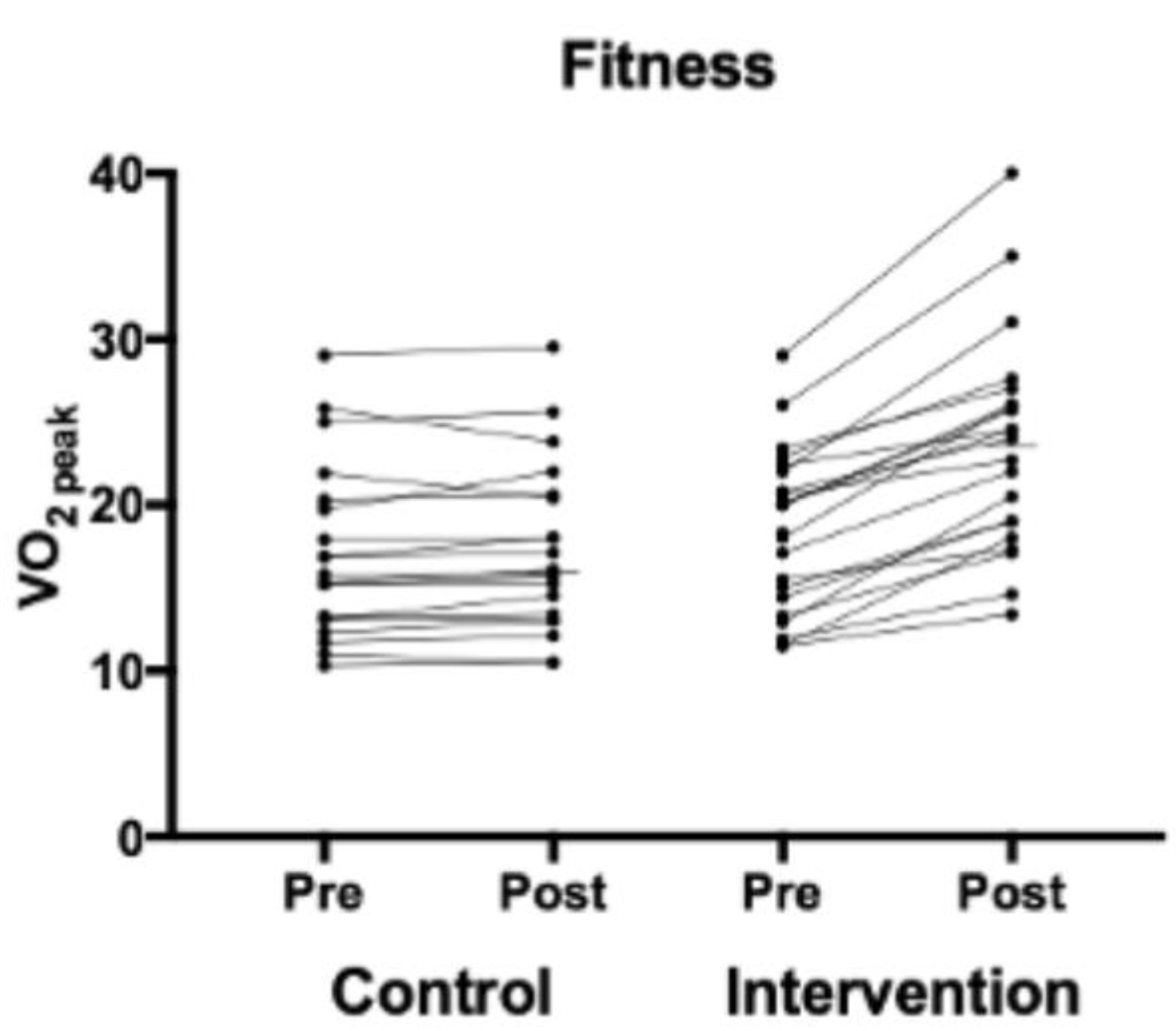
Results for cardiorespiratory fitness (VO₂peak) in the control and intervention group at pre- and post-treatment tests

**Table 1.**
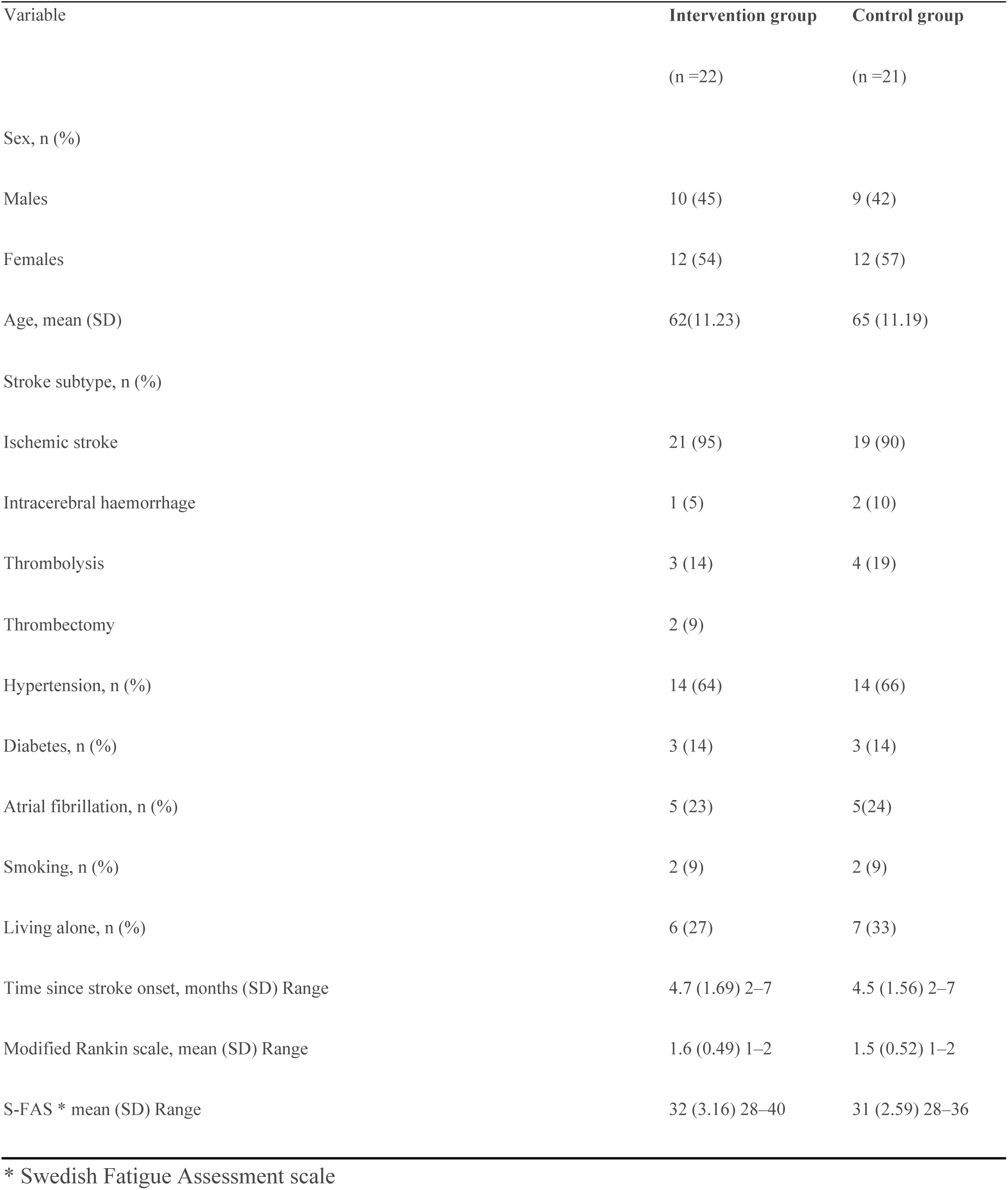
Baseline characteristics of study participants (n =43)

| Variable | Intervention group | Control group |
| --- | --- | --- |
|  | (n =22) | (n =21) |
| Sex, n (%) |  |  |
| Males | 10 (45) | 9 (42) |
| Females | 12 (54) | 12 (57) |
| Age, mean (SD) | 62(11.23) | 65 (11.19) |
| Stroke subtype, n (%) |  |  |
| Ischemic stroke | 21 (95) | 19 (90) |
| Intracerebral haemorrhage | 1 (5) | 2 (10) |
| Thrombolysis | 3 (14) | 4 (19) |
| Thrombectomy | 2 (9) |  |
| Hypertension, n (%) | 14 (64) | 14 (66) |
| Diabetes, n (%) | 3 (14) | 3 (14) |
| Atrial fibrillation, n (%) | 5 (23) | 5(24) |
| Smoking, n (%) | 2 (9) | 2 (9) |
| Living alone, n (%) | 6 (27) | 7 (33) |
| Time since stroke onset, months (SD) Range | 4.7 (1.69) 2–7 | 4.5 (1.56) 2–7 |
| Modified Rankin scale, mean (SD) Range | 1.6 (0.49) 1–2 | 1.5 (0.52) 1–2 |
| S-FAS * mean (SD) Range | 32 (3.16) 28–40 | 31 (2.59) 28–36 |

\* Swedish Fatigue Assessment scale

## Discussion

This randomized controlled trial evaluated the effects of a home-based, supervised cardiorespiratory interval training program (HS-CITP) on post-stroke fatigue (PSF) and cardiorespiratory fitness (VO₂peak) in individuals recently discharged from early supported discharge (ESD) care. The findings demonstrate that HS-CITP significantly reduced self-reported fatigue and improved cardiorespiratory fitness compared to standard ESD care alone.

The observed reduction in fatigue (mean difference: 5.3 points) was both statistically significant and clinically meaningful, particularly in light of previous meta-analyses reporting a moderate-to-strong association between cardiorespiratory fitness and fatigue (4). The significant increase in VO₂peak (+4.48mL/kg/min) further supports the efficacy of the intervention and aligns with prior evidence indicating that high-intensity exercise enhances aerobic capacity (19). Even modest gains in VO₂peak may reduce exertional fatigue and improve endurance in everyday life, given the high relative aerobic load of daily activities after stroke (8).

Importantly, qualitative findings from Svedjebrant et al. (30), derived from the same trial, enrich these quantitative outcomes. Participants described the training as safe, feasible, and a turning point in their recovery—helping them break a cycle of inactivity and fatigue. They reported increased strength, energy, and confidence in physical activity, both during the intervention and in daily life. Several participants emphasized a regained sense of control, self-efficacy, and the motivational boost of perceiving tangible improvements. These experiential accounts suggest that the observed improvements in S-FAS scores and VO₂peak were accompanied by meaningful changes in quality of life, motivation, and independence.

The intensity of training is also noteworthy. Sessions were conducted at 70%–80% of maximal heart rate and at a perceived exertion of 14–16 on the Borg scale, corresponding to high-intensity exercise. Recent reviews emphasize the superiority of high-over moderate-intensity training for improving aerobic capacity and functional outcomes in stroke survivors (31). Our findings therefore support the feasibility and benefits of prescribing high-intensity exercise relatively early after stroke.

Several physiological mechanisms may explain the observed effects, including enhanced cerebral perfusion, upregulation of neurotrophic factors such as brain-derived neurotrophic factor (BDNF), and attenuation of low-grade systemic inflammation (14, 15). The association between low cardiorespiratory fitness and more severe fatigue, particularly in individuals with blunted cardiovascular responses to exertion (12), underscores the potential of individualized aerobic training as a targeted intervention for PSF.

Our results are consistent with recent evidence showing an association between cardiorespiratory fitness and PSF. For example, a recent study reported that each 4% increase in VO₂peak per kilogram of body weight corresponded to a one-point reduction in fatigue scores (32). Together with a meta-analysis showing a significant link between physical fitness and fatigue (5), this reinforces the deconditioning hypothesis and highlights aerobic training as a promising intervention for PSF.

Strengths of this study include the high adherence rate (mean 92%), demonstrating both feasibility and acceptability of a supervised, home-based program in the early subacute phase. Consistent attendance mirrors previous findings that stroke survivors can successfully engage in structured aerobic training when provided with guidance and support (17, 20). Another strength is the use of standardized and objective measures of cardiorespiratory fitness in a vascular population. Furthermore, this was a two-centre study, which strengthens external validity and enhances generalizability of the findings.

Limitations include the modest sample size, which restricts generalizability, despite balanced baseline characteristics. The physiotherapists delivering the intervention were not blinded, which may have introduced performance bias. The recruitment period was long (2018–2025), reflecting the challenges of enrolling participants into an intervention requiring availability for three training sessions per week over eight weeks. Recruitment was further complicated by pauses during the COVID-19 pandemic, as stroke patients constitute a high-risk group. The long-term sustainability of the observed benefits also remains unknown, and future research should include extended follow-up. Moreover, baseline physical activity and training history were not assessed, which may have influenced outcomes, as participants might have been more motivated toward exercise. Larger randomized trials are needed to confirm and extend these findings.

This study has clear clinical implications. All participants received standard advice on fatigue management, but only the intervention group was offered structured, supervised aerobic training. The significant additional benefits observed indicate that advice alone is insufficient to alleviate PSF. Instead, combining education with structured exercise appears necessary to achieve meaningful improvements in both fatigue and fitness (30).

The current data support that individuals with pronounced PSF should not be excluded from such interventions. On the contrary, they may benefit most from the combination of professional support and progressive training. Given the overlapping cardiovascular risk profiles of stroke and cardiac patients, closer integration between stroke and cardiac rehabilitation could facilitate broader implementation of supervised aerobic training (16). Currently, there are no established evidence-based treatments for PSF. Our findings contribute to the growing body of evidence—paralleling results from cancer-related fatigue (33) and multiple sclerosis (34)—suggesting that exercise interventions can effectively reduce fatigue after stroke.

## Conclusions

In, summary, this study provides compelling evidence that supervised, home-based aerobic training is both effective and personally meaningful for individuals with PSF. Structured aerobic exercise may be a core element of early post-stroke rehabilitation in subjects with fatigue.

## Data Availability

All data are available without restrictions from the corresponding author on reasonable request.

## Non-standard Abbreviations and nonstandard acronyms

ESD: Early Supported Discharge
HIIT: High-intensity interval training
HS-CITP: Home-based, supervised cardiorespiratory interval training program
LIVI: Lugnet Sports Science Institute laboratory
PSF: Post-stroke Fatigue
RPE: Borg Rating of Perceived Exertion scale
S-FAS: Swedish Fatigue Assessment Scale
UMEX: Umeå Movement and Exercise Laboratory
VO₂peak: Peak oxygen consumption

## Acknowledgments

We thank registered physiotherapist, PhD, Tobias Stenlund for carrying out the incremental cardiopulmonary exercise test at the UMEX laboratory at Umeå University and registered biomedical scientist Lars Wedholm at the Lugnet Sports Science Institute (LIVI) laboratory, Dalarna University in Falun. We also thank research nurse, Mariann Haapalahti at Strokecenter, the University hospital in Umeå for handling pre- and post-assessment of the S-FAS, and PhD statistician, Per Liv, Department of Public Health and Clinical Medicine, for support with the analyses.

